# Nav1.7 and Nav1.8 form supramolecular active clusters with TRKB in mouse and human DRG neurons during development of neuropathic pain

**DOI:** 10.1101/2022.11.05.22281929

**Authors:** Liting Sun, Hang Xian, Yunxin Shi, Taotan Yang, Hongyan Shuai, Ruilong Xia, Ting Wen, Wei Xia, Ran Qian, Fengting Zhu, Yuanying Liu, Zhicheng Tian, Lamei Li, Rui Cong, Ceng Luo, Shengxi Wu, Xiafeng Shen, Xin Yu, Rou-Gang Xie, Changgeng Peng

## Abstract

Neuropathic pain affects 7-10% of the global population, and one of its characteristics is sensitization of somatosensory nervous system. Altered expression of ion channels and receptors has been found to be involved in neuronal hyperexcitability after injury to somatosensory nervous system, it is, however, unknown that if ion channels and receptors could gain qualitative changes on the level of structure organization when they are excessively expressed in same one neuron during the development of neuropathic pain. Here we show first that not only the expression of voltage-gated sodium channels Nav1.7 (SCN9A), Nav1.8 (SCN10A) and TRKB (also named NTRK2) increased in DRG neurons of patients with over 3-month severe neuropathic pain induced by brachial plexus avulsion (BPA), but also Nav1.7 and Nav1.8 formed supramolecular active clusters with or without TRKB in DRG neurons of mice with chronic neuropathic pain induced by spared nerve injury or diabetic neuropathy and of BPA pain patients with neuropathic pain. Nav1.7, Nav1.8 and TRKB might function in a coordinated manner in orderly organized supramolecular active clusters to geometrically increase the hyperexcitability of pathological DRG neurons. Our findings suggest that supramolecular active clusters of Nav1.7, Nav1.8 and TRKB might need be targeted for curing neuropathic pain, and that inhibition of both Nav1.7 and Nav1.8 might be required to achieve efficient relief of neuropathic pain.

## Introduction

Neuropathic pain affects 7-10% of the global population (*1, 2*), but is lack of efficient drugs (*3*). Voltage-gated sodium channels (VGSCs) are potential targets for pain relief. The first-generation unspecific VGSCs inhibitors lidocaine and mexiletine are local anesthetics used in clinic, but they have side effects due to the inhibition of certain VGSC alpha-subtypes in the brain (e.g. Nav1.1 and Nav1.2) and heart (e.g. Nav1.5) (*4*). Therefore the idea to develop second-generation sodium channel blockers with Nav subtype selectivity was raised. Nav1.8 (SCN10A, also named PN3 and SNS), a member of VGSC family, was first identified to be a Tetrodotoxin-resistant sodium channel and primarily expressed in small neurons of rat DRG in 1996 by A. N. Akopian, et al. and L. Sangameswaran et al. (*5, 6*), and the mouse *Scn10a* cDNA was cloned by Souslova et. Al., in 1997 (*7*). Functional study showed that Nav1.8 knock-out mice have an elevated mechanical pain threshold to noxious pressure and also have deficits in inflammatory and visceral pain, but not neuropathic pain (*8, 9*). These findings suggested that inhibition of Nav1.8 may produce analgesia. In line with phenotype of loss-of-function, gain function of Nav1.8 mutations was found in patients with painful small-fiber neuropathy or with lower mechanical pain sensitivity, and patch-clamp experiments demonstrated that the Nav1.8 mutation enhanced the channel’s response to depolarization and produce hyperexcitability in DRG neurons (*10-13*). These findings together supported that Nav1.8 is a potential target for development of second-generation of sodium channel blockers to treat acute pain.

Nav1.7 (SCN9A, also named PN1), another member of VGSCs, was originally found in mouse in 1996 by M. C. Beckers et al. and C. A. Kozak, et al. (*14, 15*). The Nav1.7 cDNA was cloned from rat PC12 cells in 1997 and found to be principally expressed in peripheral neurons (*16, 17*). In 2000, Lai J et. al. reported that knocking down the expression of Nav1.7 using *Scn9a* antisense in DRG neurons alleviated neuropathic pain (*18*). Later on it’s found that gain-of-function mutations of SCN9A caused inherited erythromelalgia and idiopathic small fiber neuropathies (*19-21*). Nassar et. al. reported that the mice with conditional knock-out *Scn9a* in Nav1.8 neurons using a *Nav1*.*8-Cre* had attenuated acute and inflammatory pain, but normal response to heat-evoked pain (*22*). The depletion of Nav1.7 in all DRG neurons of mice using *Advillin-Cre* abolished mechanical pain, inflammatory pain and certain type of heat pain, but neuropathic pain were not affected, and the further ablation of Nav1.7 in both sensory and sympathetic neurons eliminated all kinds of pain (*23*). More importantly, the loss of function mutation of SCN9A leads to congenital insensitivity to pain in human (*24*). These findings together suggest that Nav1.7 is a potential target for the treatment of acute pain and neuropathic pain.

Given the strong clinical relevance of Nav1.7 and Nav1.8 in neuropathic pain, a second-generation sodium channel blockers selectively targeting Nav1.7 or Nav1.8 have been developed since as early as 2009, but none of them has achieved efficient effect in attenuation of neuropathic pain in clinical trials yet. This raised the possibility that Nav1.7 and Nav1.8 could happen some critical changes during the development of neuropathic pain. Fully unveiling the changes of Nav1.7 and Nav1.8 in pathological condition could help to develop VGSC selective blocker drugs for neuropathic pain treatment.

We previously found that Nav1.7 and Nav1.8 are simultaneously regulated by *miR-96* and the depletion of *miR-96* in mice leads to mechanical and heat allodynia (*25*). Here we show that both the expression of Nav1.7 in DRG neurons and the expression of Nav1.8 in Nav1.7-expressing neurons increased in mice with neuropathic pain induced by spared nerve injury (SNI) and in patients with neuropathic pain induced by brachial plexus avulsion (BPA). Moreover, we found that Nav1.7 and Nav1.8 formed orderly organized supramolecular active cluster in DRG sensory neurons of mice with chronic neuropathic pain induced by SNI or Strepotozocin and in BPA patients suffering from severe chronic neuropathic pain. The qualitative change of Nav1.7 and Nav1.8 on the level of structure might contribute to the hyperexcitability of DRG neurons.

## Materials and methods

### Animals

In this study, all animals were housed 5 per cage, at 21°C, 50% humidity, on a 12h light:12h dark schedule in the SPF standard animal facility in accordance with the guidelines of Tongji University and all animal work was conducted under ethical permission from Tongji University ethical review panel.

Surgery: Spared nerve injury (SNI) surgical procedures were performed under isoflurane-induced anesthesia as previously described (*26, 27*). Briefly, a small incision was made below the left biceps femoris followed by blunt dissection of the muscle to expose three terminal branches of the sciatic nerve: common peroneal, tibial and sural nerves; The common peroneal and tibial nerves were tightly ligated with 6-0 silk suture, transected together distally to the ligation and a piece of 1-2 mm of the nerves was removed from the distal stump avoiding any contact with or stretching of the intact sural nerve during the surgery process. Finally, skin was closed with 5-0 Nylon sutures. Mechanical threshold was measured before and at 2 weeks and 6 weeks post operation.

Diabetic mouse model: Wild type C57BL/6 mice at age of 4-5 weeks were intraperitoneally injected 55 mg/kg Streptozotocin (STZ, dissolved in 100 mmol/L citrate buffer, pH 4.5) after 12 hours food-fasting and 2 hours water deprivation, followed by 4 consecutive intraperitoneal injection of 55 mg/kg STZ after 6 hours food-fasting once a day, and 2 hours water deprivation after each injection. Two weeks post last injection of STZ, blood glucose was measured after 6 hours food-fasting. Mice with fasting blood glucose more than 13 mM were designated to diabetic group. Mechanical threshold of diabetic mice was measured at 4 weeks and 10 weeks post last injection of STZ. Only the diabetic mice which have mechanical threshold⩽0.4 g at 4 weeks were used for the later experiment.

### Behavioral test

Before behavioral test, mice were placed in transparent Plexiglass chambers on top of a raised wire mesh and allowed to acclimate for 20 minutes.

Mechanical threshold: The paw withdrawal threshold of the hind paws was measured with a set of calibrated monofilaments (von Frey hairs, DanMic Global, USA) in order of increasing forces from 0.008 g to 2 g. Each monofilament was applied five times. The force at which the animal withdrew the paw from at least three out of five stimuli was recorded as paw withdrawal threshold.

### Human tissues

Patients with brachial plexus avulsion (BPA) often suffered from severe neuropathic pain in the affected extremity, in addition to paralysis of muscles after injury. A series of reconstruction surgery can help BPA patients to restore movement of the ipsilateral arm and partially alleviate pain (*28*). The avulsed roots of the affected brachial plexus and the dislocated cervical DRG tissues were resected during the surgical process of nerve reconstruction or transplantation under the ethic permit (KY20222228) approved by the Ethics Committee of Xijing Hospital, the first affiliated hospital of Fourth Military Medical University. Normal cervical DRG tissues from naturally aborted 29-weeks human embryos were collected according to the ethic permit (MECDU-201909-1) approved by Dali University. The collected human tissues were then either lysed with RIPA buffer containing protease inhibitors, or immersed in 4% paraformaldehyde for 24∼48 h and then in 30% sucrose for 24 h at 4℃, and embedded in OCT, further sectioned in thickness of 12 μm using a cryostat.

### Immunostainings

Mouse DRGs were processed in a same way as human DRG did, and sectioned in thickness of 14 μm. Immunostainings were performed as previously described (*29*). Briefly, sections were incubated with a blocking solution containing 0.2% Triton X-100 and 5% bovine serum albumin (BSA) for 1 h at room temperature, followed by incubation of primary antibodies overnight at 4 °C, and then sections were washed three times with PBS, incubated with the secondary antibodies for 2 h at room temperature. After washed three times with PBS, sections were mounted with anti-fluorescence quenching solution and coverslip. Fluorescent images were captured using Olympus confocal microscope (Olympus FV3000, Ishikawa, Japan) or Zeiss confocal microscope (LSM880, LSM780, Carl Zeiss, Oberkochen, Germany) or fluorescence microscope (Vert.A1, Carl Zeiss, Oberkochen, Germany) and processed with Adobe Photoshop software.

The following primary antibodies were used: mouse antibodies against SCN10A (NeuroMab, 75-166, 1:200) and NeuN (Merck, MAB377, 1:100), rabbit antibodies against SCN9A (Proteintech Group, 20257-1-AP, 1:200) and NeuN (Cell Signaling Technology, 24307S, 1:200), and goat antibody against TrkB (R&D system, AF1494, 1:400). Secondary antibodies were conjugated with AlexaFluor 488/555/647 (Thermo Fisher Scientific, Massachusetts, USA, 1:1000).

### Quantification methods

SCN9A, SCN10A, TrkB and NeuN positive cells were counted on 6 every ninth serial sections from L4-L5 DRG of Sham and SNI mice (n=3-6) at 2 weeks or 6 weeks post operation or on 4-6 sections of human cervical DRGs (n=3-4) using Image J.

## Statistical analysis

All numerical data were shown as individual raw data points with mean and standard deviation (mean ± SD) and analyzed by GraphPad Prism 6 software (GraphPad Software Inc, CA, USA) The D’agostino-Pearson omnibus normality test was performed before further analysis. The non-parametric Kruskal-Wallis test was used when the data were not normally distributed. The numerical data of immunostaining results between 2 groups like Sham and SNI mice were analyzed by unpaired Student’s T-test. A value of *p* < 0.05 was considered as statistically significant.

## Results

### Dynamic changes of the number of SCN9A+/SCN10A+ DRG neurons in mice after nerve injury

Since the expression of SCN9A and SCN10A is co-regulated by *miR-96* and *miR-96* is down-regulated in DRG after nerve injury, it’s possible that the expression of SCN9A and SCN10A is up-regulated in same one (type) DRG neurons after nerve injury. We deeply investigated the expression of SCN9A and SCN10A in mouse DRG from 2 weeks to 6 weeks post operation (WPO) of SNI. The percentages of SCN9A^+^ and SCN9A^+^/SCN10A^+^ neurons in L4 DRG increased significantly in SNI mice at 2 WPO, and there was also a trend in increase of SCN10A^+^ neurons in SNI mice at 2 WPO when compared to that in Sham mice (Fig. 1A, B). However, by 6 WPO the percentage of SCN9A^+^/SCN10A^+^ neurons significantly decreased in L4 DRG (Fig. 1A, B). Compared to Sham mice, the percentages of SCN9A^+^ neurons significantly decreased in L5 DRG of SNI mice at 2 WPO, and the percentages of SCN9A^+^/SCN10A^+^ neurons dropped in L5 DRG of SNI mice at both 2 and 6 WPO (Fig. 1C, D). Further analysis showed that the proportion of SCN9A^+^/SCN10A^+^ neurons to total SCN9A^+^ neurons was neither changed in L4 and L5 DRG of SNI mice at 2 WPO, nor in L4 DRG at 6 WPO, but significantly decreased in L5 DRG of SNI mice at 6 WPO (Fig. 1E) when compared to that in Sham mice. The proportion of SCN9A^+^/SCN10A^+^ neurons to total SCN10A^+^ neurons significantly increased in L4 DRG of SNI mice at 2 WPO when compared to that in Sham mice, not in L4 DRG at 6 WPO, and not in L5 DRG of SNI mice at both 2 and 6 WPO (Fig. 1F). These results suggested that the part of increased SCN9A^+^/SCN10A^+^ neurons in L4 DRG of SNI mice at 2 WPO was from original SCN10A^+^/SCN9A^-^ neurons that gained expression of SCN9A after nerve injury, however it couldn’t rule out that some of neurons expressing SCN9A^+^/SCN10A^+^ may come from original SCN10A^-^/SCN9A^+^ and/or SCN10A^-^/SCN9A^-^ neurons.

**Fig. 1.**
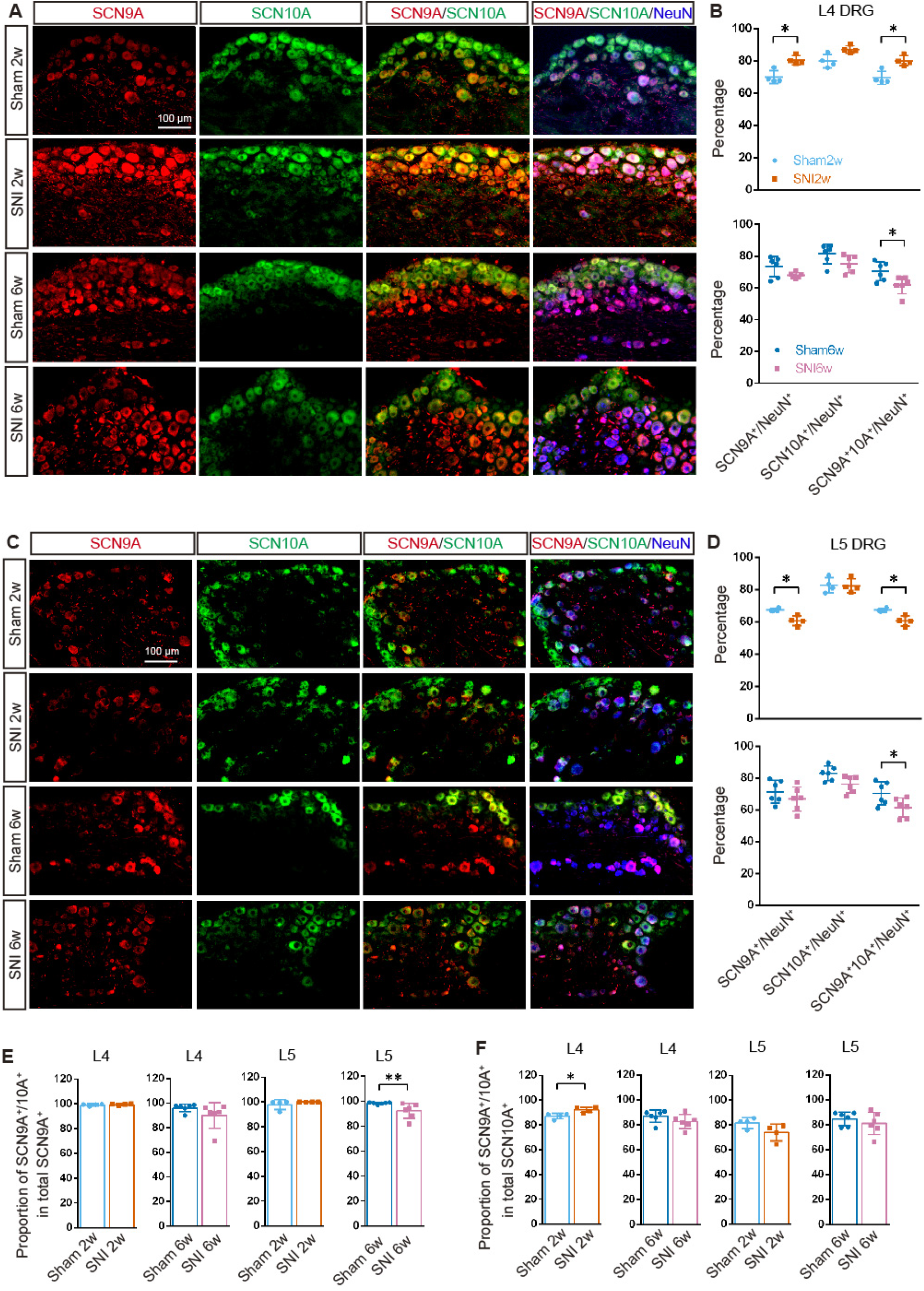
Dynamic changes of the numbers of SCN9A^+^ and SCN10A^+^ neurons in L4 and L5 DRGs of SNI mice at 2 weeks or 6 weeks post operation (WPO). (A-B) Quantitative analysis of the immunostained L4 DRG sections with antibodies against SCN9A (red), SCN10A (green) and NeuN (blue) showed that the percentages of both SCN9A^+^ and SCN9A^+^/SCN10A^+^ neurons in L4 DRG of SNI mice significantly increased at 2 WPO when compared to that in Sham mice, and the percentage of SCN9A^+^/SCN10A^+^ neurons in L4 DRG of SNI mice significantly decreased at 6 WPO when compared to that in Sham mice (B). (C, D) The percentage of SCN9A^+^/SCN10A^+^ neurons in L5 DRG of SNI mice significantly decreased at 2 and 6 WPO when compared to that in Sham mice, and that the percentage of SCN9A^+^ neurons in L5 DRG of SNI mice significantly decreased at 2 WPO when compared to that in Sham mice. (E) The proportions of SCN9A^+^/SCN10A^+^ neurons among total SCN9A^+^ neurons did not statistical significantly change in L4 DRG of SNI mice either at 2 WPO or 6 WPO when compared to that in Sham mice, but it’s decreased significantly in L5 DRG of SNI mice at 6 WPO when compared to that in Sham mice. (F) The proportion of SCN9A^+^/SCN10A^+^ neurons among total SCN10A^+^ neurons statistical significantly increased only in L4 DRG of SNI mice at 2 WPO, not at 6 WPO when compared to that in Sham mice. Scale bar=100 μm, * p<0.05, ** p<0.01, unpaired Student’s T-test, n=4-6.

### Formation of Supramolecular active clusters of Nav1.7/Nav1.8/TrkB in DRG neurons of SNI mice

Since increase of SCN9A and SCN10A in the same one neuron could relate to hyperexcitability of DRG neuron in *miR-96* knock-out mice which have mechanical and heat allodynia, we carefully observed SCN9A^+^/SCN10A^+^ neurons and found that SCN9A, SCN10A and TrkB formed clusters on and along plasma membrane in DRG neurons of SNI mice at 2 WPO (Fig. 2A). Small clusters of SCN9A/SCN10A/TrkB were also found in near to the membrane of DRG neurons of Sham mice at 2 WPO (Fig. 2A). By 6 WPO clusters of SCN9A/SCN10A/TrkB grew into supramolecular active clusters (SMACs, active domain) in SNI mice, but not in Sham mice (Fig. 2A). Quantitative analysis showed that the number of cells containing cluster of SCN9A/SCN10A dramatically increased in DRG of SNI mice compared to that in Sham mice at both 2 and 6 WPO (Fig. 2B), the majority of these SCN9A/SCN10A cluster-containing cells was TrkB^+^ neurons (Fig. 2C, D), and there was a dramatically increase of SCN9A/SCN10A cluster-containing TrkB^+^ neurons in SNI mice compared to that in Sham mice from 2 to 6 WPO (Fig. 2C). Cluster of SCN9A/SCN10A was also formed in more TrkB^-^ neurons in SNI mice than in Sham mice, with statistical significance at 6 WPO (Fig. 2D). The average diameter of SCN9A particles in clusters was significant larger in DRG neurons of SNI mice at 6 WPO than that in Sham mice (Fig. 2E), and the diameters of SCN10A particles and TrkB particles in clusters were also significantly bigger in DRG neurons of SNI mice from 2 to 6 WPO compared to that in Sham mice (Fig. 2F, G). The size of particles of SCN9A, SCN10A and TRKB in DRG neurons of SNI mice significantly enlarged from 2 to 6 WPO (Fig. 2A, E-G).

**Fig. 2.**
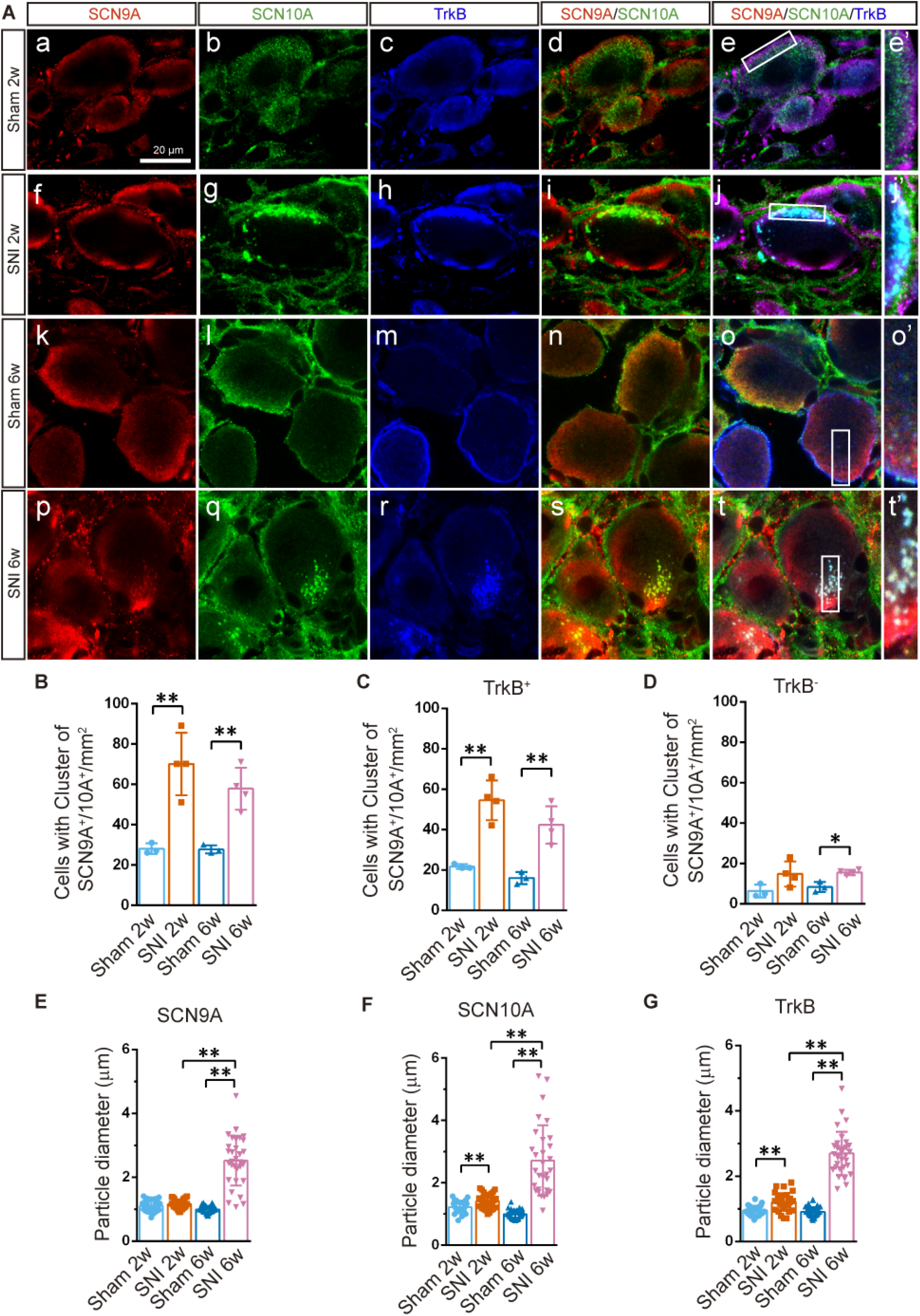
Clusters of SCN9A and SCN10A appeared in L4-5 DRGs of SNI mice at 2 WPO and 6 WPO. (A) Immunostaining of SCN9A (red), SCN10A (green) and TrkB (blue) on L4-L5 DRG sections from Sham (a-e) and SNI (f-j) mice at 2 WPO, as well as from Sham (k-o) and SNI (p-t) mice at 6 WPO showed that cluster of SCN9A/SCN10A/TrkB formed in DRG from SNI mice at 2 WPO (f-j), and developed into supramolecular cluster by 6 WPO (p-t), e’, j’, o’ and t’ are the high magnification view of inset in e, j, o, t, respectively. Scale bar=100 μm. (B) The numbers of cells containing cluster of SCN9A/SCN10A were increased in L4-5 DRG of SNI mice at both 2 and 6 WPO when compared to that in Sham mice. (C) The numbers of cells contained cluster of SCN9A/SCN10A/TrkB were increased in L4-5 DRG of SNI mice at both 2 and 6 WPO when compared to that in Sham mice. (D) SNI mice had more TrkB^-^ cells contained cluster of SCN9A/SCN10A than Sham mice. (E) The diameters of SCN9A particles in cluster of DRG neurons were significantly bigger in SNI mice than that in Sham mice at 6 WPO. (F) The diameters of SCN10A particles in cluster of DRG neurons were significantly bigger in SNI mice than that in Sham mice at both 2 and 6 WPO. (G) The diameters of TrkB particles in cluster of DRG neurons were significantly bigger in SNI mice than that in Sham mice at both 2 and 6 WPO. * p<0.05, ** p<0.01, unpaired Student’s T-test, n=3-4.

### Increased population of SCN9A/SCN10A/TRKB DRG neurons in patients with BPA

To investigate if human DRG has a similar change as mouse DRG after nerve injury, immunostaining of TRKB, SCN9A and SCN10A on sections of pathological DRG from BPA patients with server neuropathic pain over 3-month and of normal DRG from naturally aborted 29-weeks embryos was carried out. It obviously showed that there were some TRKB/SCN9A neurons expressed no or rare level of SCN10A in normal DRG (Fig. 3 A-D), in contrast, most of TRKB/SCN9A neurons in pathological DRG highly expressed SCN10A (Fig. 3 E-H). To examine if the proportion of SCN9A, SCN10A and TRKB neurons was changed after nerve injury, immunostaining analysis was carried out for SCN9A/TRKB/NEUN, or SCN10A/TRKB/NEUN, or SCN9A/SCN10A/TRKB on sections of pathological DRG from BPA patients and of normal DRG from 29-weeks abortive embryos (Fig. 4A-C). Quantitative analysis of immunostaining reaction positive neurons showed that the numbers of neurons with high expression of SCN9A, SCN10A and TRKB significantly increased, and the neurons with low expression of SCN9A, SCN10A and TRKB decreased, in pathological DRG compared to that in normal DRG (Fig. 4D). The proportion of SCN9A/SCN10A/TRKB neurons was significantly elevated in the pathological DRG than that in normal DRG (Fig. 4E). Further analysis showed more TRKB neurons expressing SCN9A in pathological DRG than that in normal DRG (Fig. 4F), especially, within the type of TRKB neurons the proportion of cells expressing high level of SCN9A or SCN10A remarkably increased in pathological DRG compared to that in normal DRG (Fig. 4G). Moreover, the number of TRKB/SCN9A neurons expressing high level of SCN10A (SCN10A^High+^) dramatically increased and the number of TRKB/SCN9A neurons expressing low level of SCN10A (SCN10A^Low+^) decreased in pathological DRG when compared to that in normal DRG (Fig. 4H).

### SMACs of SCN9A/SCN10A/TRKB developed in DRG neurons of BPA patients with neuropathic pain

As in mouse, SCN9A, SCN10A and TRKB formed very big SMACs in most of DRG neurons of BPA patients with chronic severe pain, but no cluster of SCN9A/SCN10A/TRKB found in control normal DRG (Fig. 5A, B). These SMACs contained elegantly organized particles which of most were constituted by SCN9A/SCN10A/TRKB, few particles containing only SCN10A or TRKB (Fig. 5B, C, Fig. S1). The diameters of particles of SCN9A, SCN10A and TRKB within SMACs ranged from 1.1-5.8 μm, with an average size at about 2 μm (Fig. 5D). It’s also found that SCN9A/SCN10A clusters were widely distributed in the nerve of DRG from BPA patients with neuropathic pain, and that these clusters contained SCN9A/SCN10A co-localized particles (Fig. 5E).

**Fig. 3.**
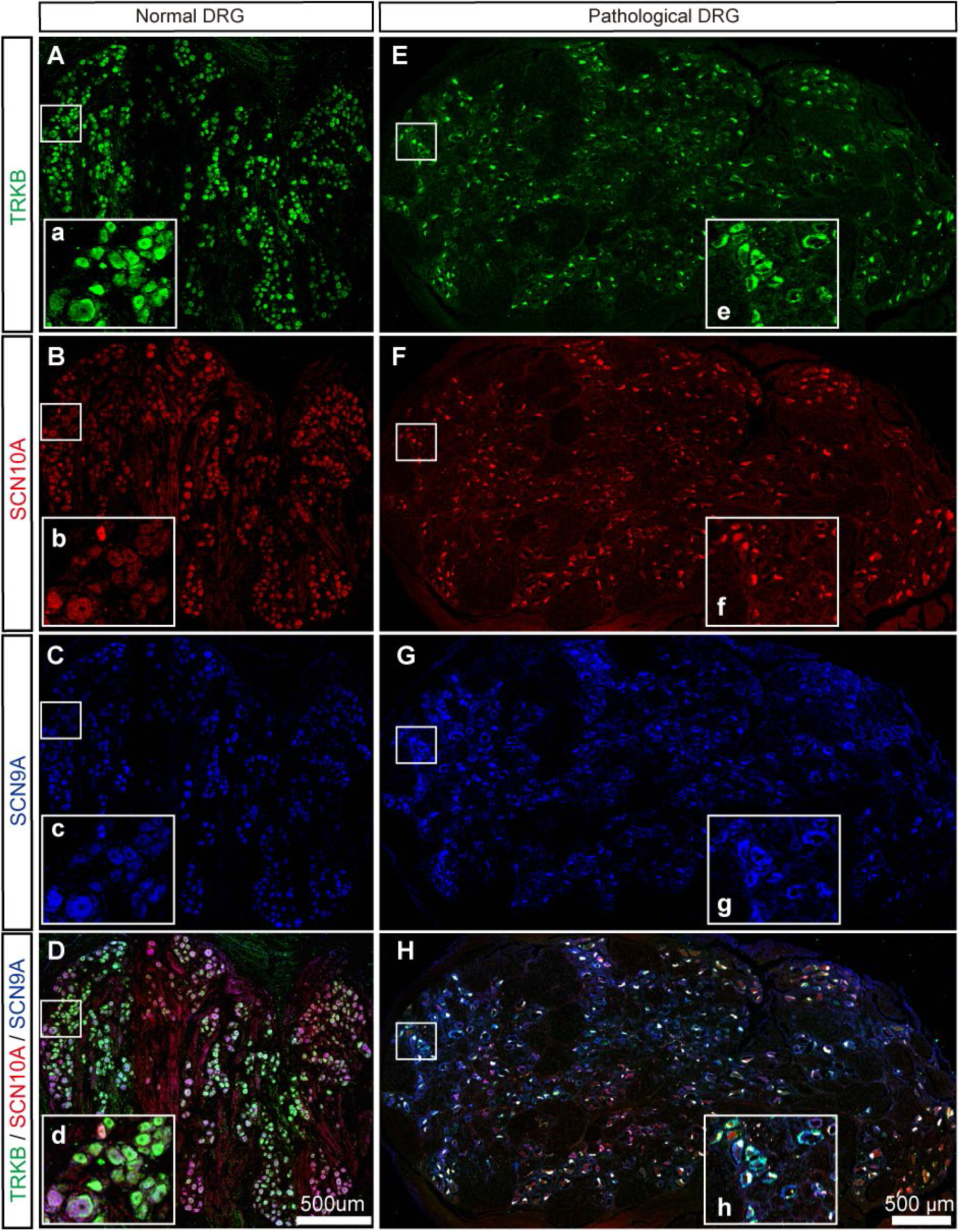
Elevated expression of SCN10A in SCN9A+/TRKB+ neurons in DRG of BPA patients. (A-H) Immunostaining of SCN9A (blue), SCN10A (red) and TRKB (green) showed that compared to normal DRG from 29-weeks abortive embryos (A-D), pathological DRG of BPA patients had more TRKB/SCN9A neurons expressing high level of SCN10A (E-H), a-h are high magnification view of inset in A-H, Scale bar=500 μm.

**Fig. 4.**
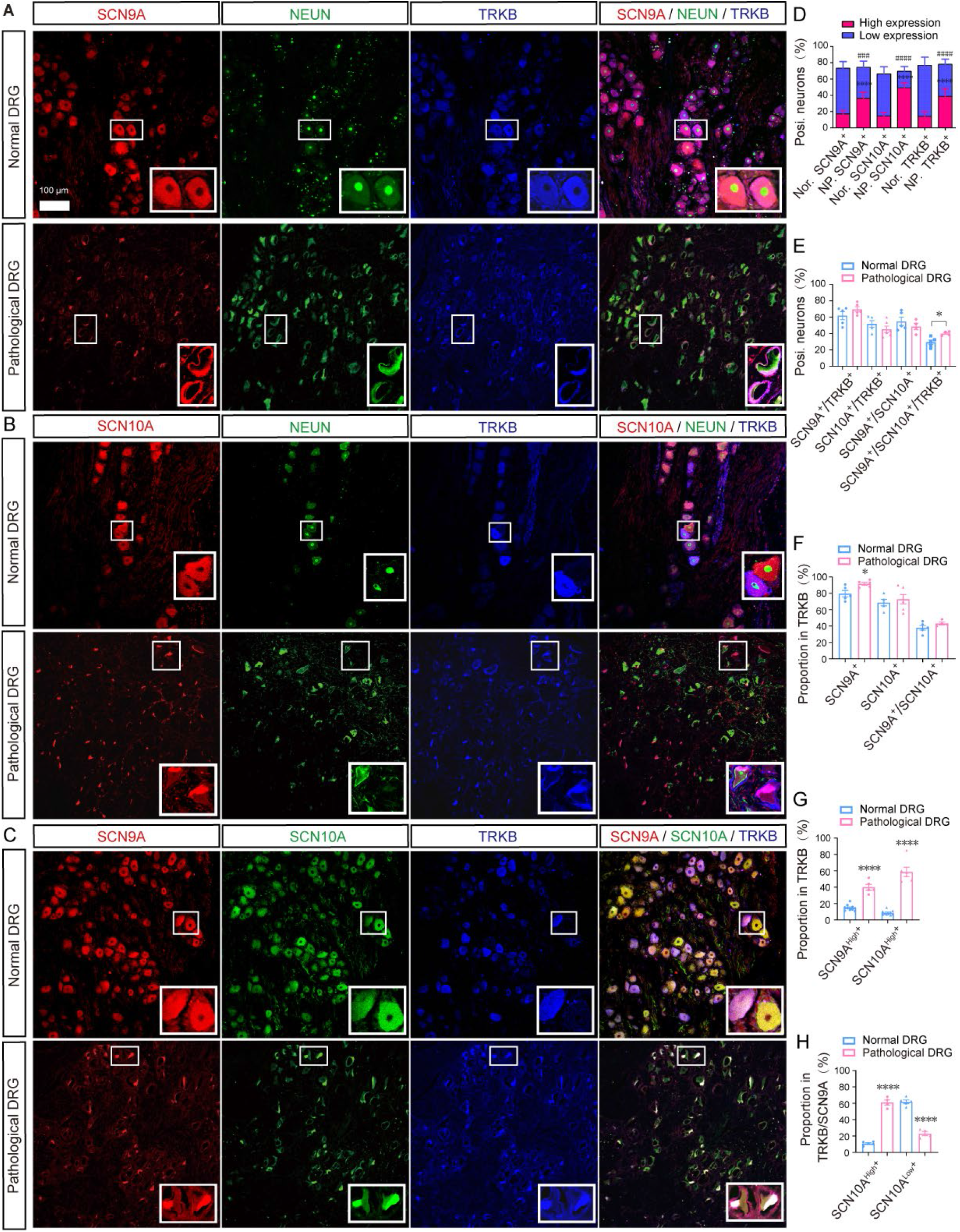
Upregulated expression of SCN9A, SCN10A and TRKB in human pathological DRG. (A-C) Immunostaining of SCN9A/TRKB/NEUN (A), or SCN10A/TRKB/NEUN (B), or SCN9A/SCN10A/TRKB (C) on sections of normal DRG from 29-weeks abortive embryos (upper panels in A-C) and of pathological DRG from BPA patients (lower panels in A-C). (D) The proportion of high (bar in red) and low (bar in blue) expression of SCN9A, SCN10A and TRKB neurons in normal DRG and pathological DRG, # represented the comparison of low expression in two groups, * represented the comparison of high expression in two groups. (E) The proportion of SCN9A/TRKB, SCN10A/TRKB and SCN9A/SCN10A/TRKB neurons in normal DRG and pathological DRG. (F) The percentage of SCN9A, SCN10A and SCN9A/SCN10A within TRKB neuronal population. (G) The proportion of neurons expressing high level of SCN9A (SCN9A^High+^) or SCN10A (SCN10A^High+^) within TRKB neuronal population. (H) The proportion of neurons expressing high level of SCN10A (SCN10A^High+^) within TRKB/SCN9A sub neuronal population. Scale bar=100 μm. * p<0.05, *** p<0.001, **** p<0.0001, ### p<0.001, #### p<0.0001, unpaired Student’s T-test, n=3-4.

**Fig. 5.**
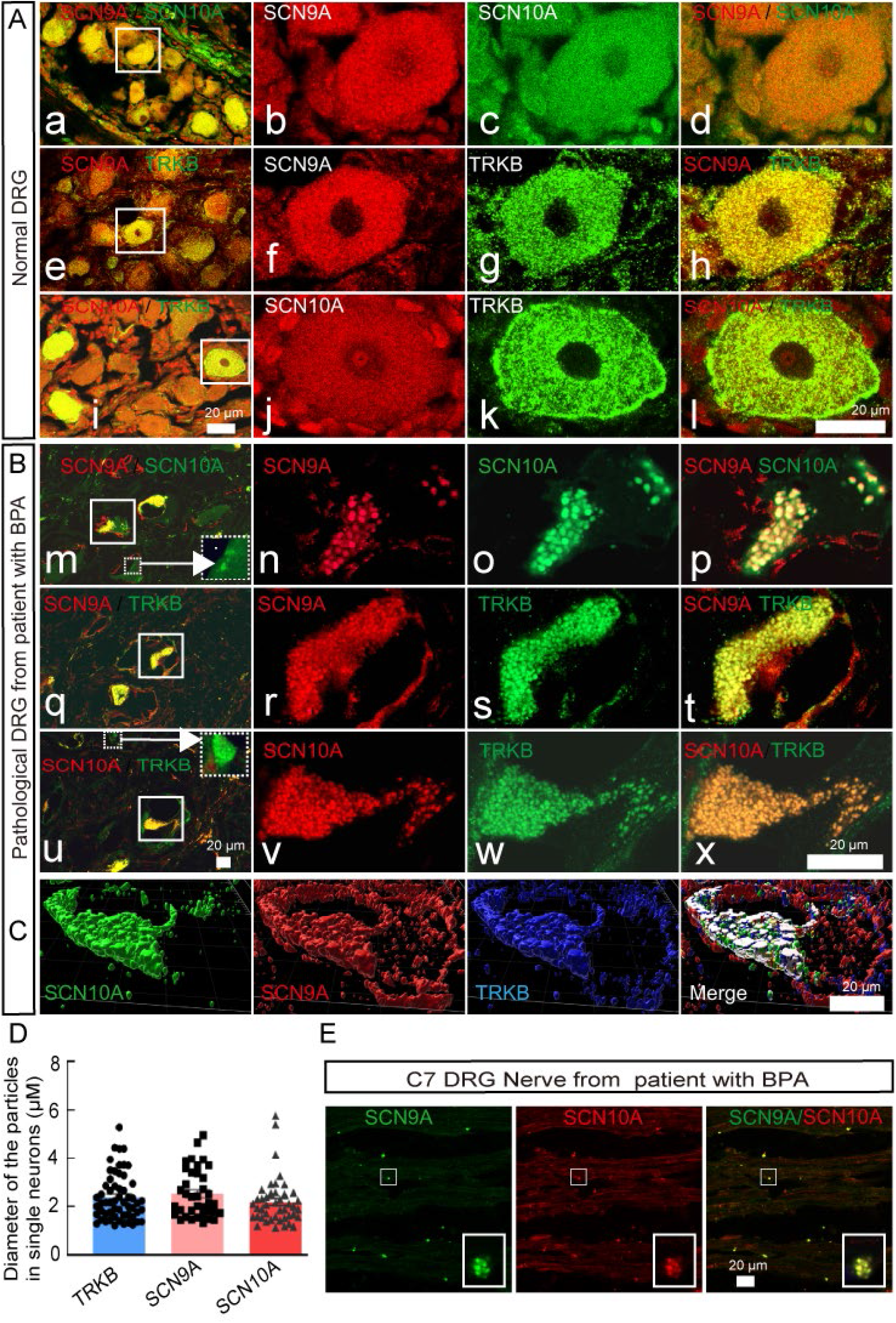
Supramolecular cluster of SCN9A/SCN10A/TRKB developed in DRG neurons of BPA patients. (A-B) Immunostaining for SCN9A/SCN10A (a-d, m-p) or SCN9A/TRKB (e-h, q-t) or SCN10A/TRKB (i-l, u-x) on sections from normal DRG (a-l) of 29-weeks abortive embryos and pathological DRG (m-x) of BPA patients showed that supramolecular clusters of SCN9A/SCN10A/TRKB on and along cytoplasm membrane of DRG neurons were only detected in pathological DRG neurons (B), not in normal DRG (A). (C) Three-dimension view of SCN9A/SCN10A/TRKB supramolecular cluster in pathological DRG neuron. (D) The diameters of the particles of TRKB, SCN9A and SCN10A in single neuron from patients with BPA. (E) Immunostaining of SCN9A (green) and SCN10A (red) showed SCN9A/SCN10A clusters in pathological DRG nerve from BPA patients. Arrows pointed to small particles containing only SCN10A or only TrkB. Scale bar=100 μm.

### Elevated expression of SCN9A, SCN10A and TRKB in DRG of BPA patients

To examine if the expression levels of SCN9A, SCN10A and TRKB increased in pathological DRG from BPA patients, we employed Western-blot and found that the expression levels of SCN9A, SCN10A and TRKB in pathological DRG elevated 1.3-fold, 1.6-fold and 1.6-fold, respectively, when compared to that in normal control DRG from 29-weeks abortive embryos (Fig. 6).

**Fig. 6.**
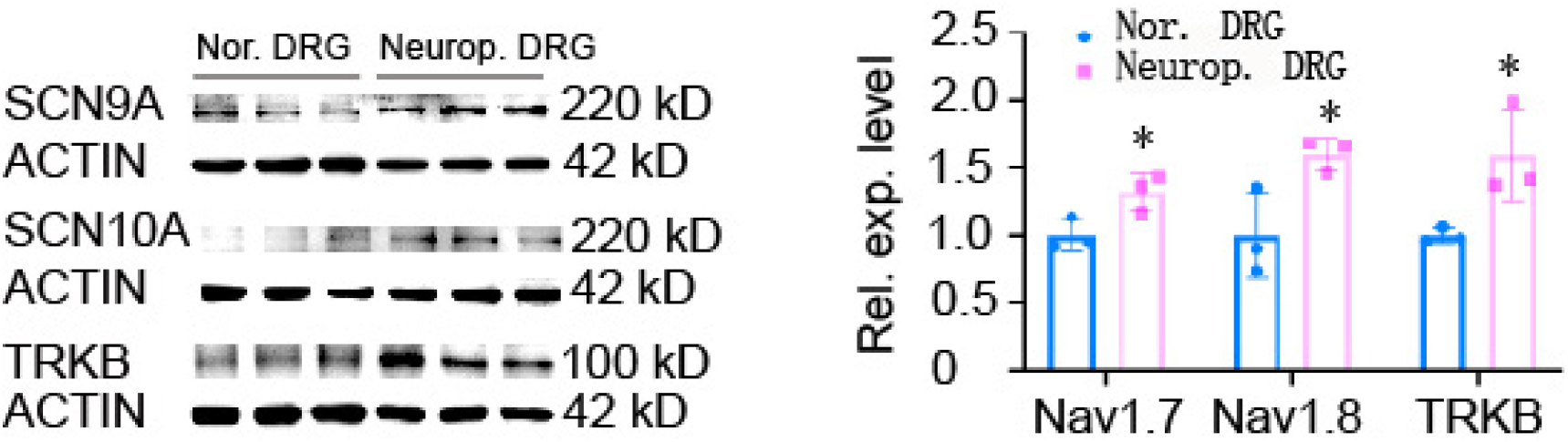
Upregulated expression of SCN9A, SCN10A and TRKB in pathological DRG from BPA patient. Western-blot results demonstrated that the expression of SCN9A, SCN10A and TRKB significantly increased in DRGs from BPA patients when compared to normal DRGs from 29-weeks abortive embryos. * p<0.05, unpaired Student’s T-test, n=3.

### SMACs of SCN9A/SCN10A formed in DRG neurons of mice with diabetic neuropathic pain

Diabetic neuropathic pain is one of common neuropathic pain. We wonder if cluster of SCN9A/SCN10A is also involved in developing of chronic neuropathic pain induced by diabetes. Diabetic C57BL/6 mouse model was constructed by intraperitoneally injection of streptozocin (STZ) and finalized with a criteria of fasting blood glucose concentration (i.e. > 13 mM at 2 weeks post last STZ-injection (WPLI)) (Fig. 7A, B). The diabetic mice which developed mechanical pain (i.e. ⩽0.4 g) at 4 WPLI were kept for later experiment, and their mechanical threshold maintained at lower level till 10 WPLI (Fig. 7C). Immunostaining of SCN9A and SCN10A demonstrated that DRG neurons of STZ-induced diabetic mice at 10 WPLI had large SMACs of SCN9A/SCN10A which were not found in the control DRG from naïve mice (Fig. 7D).

**Fig. 7.**
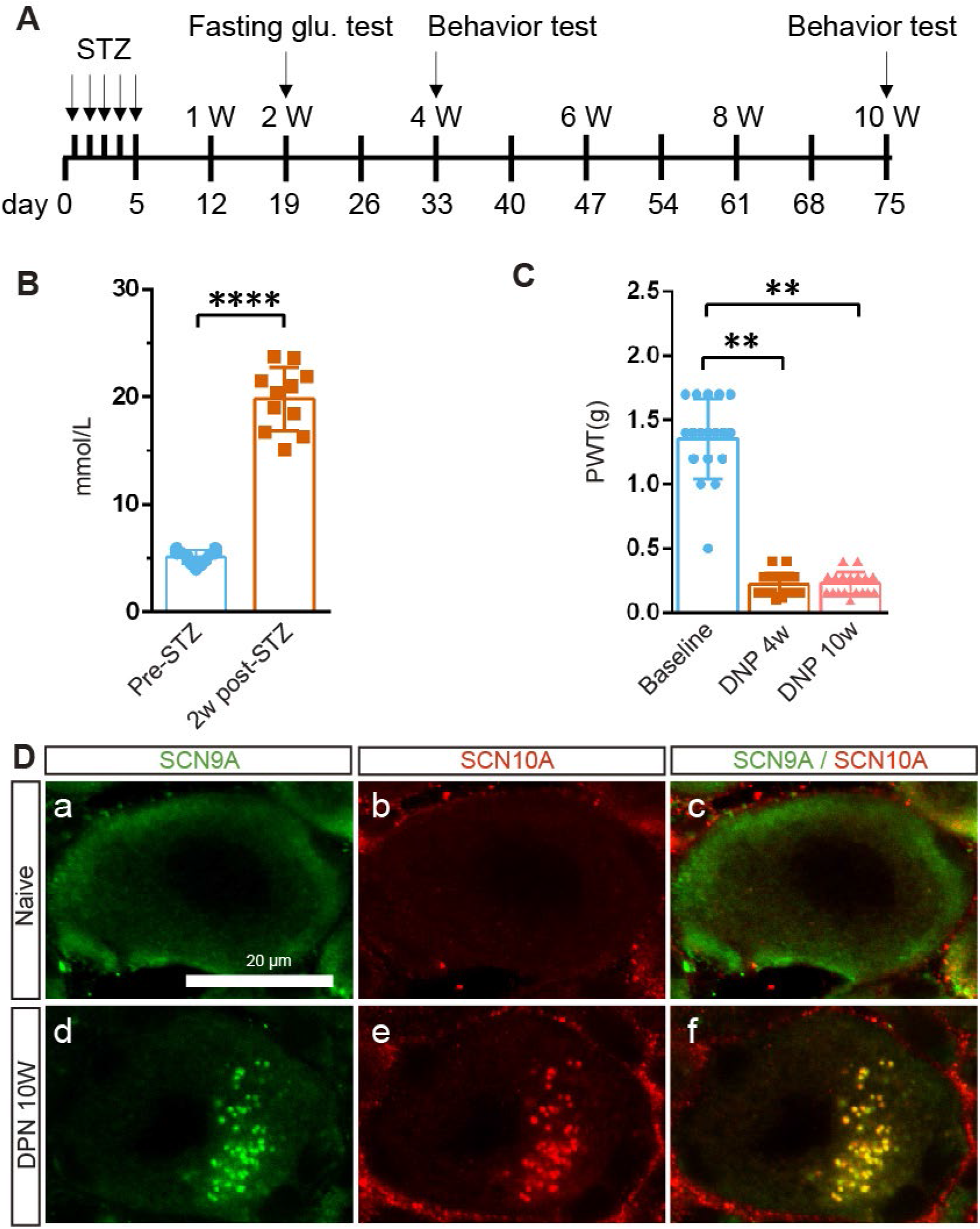
Supramolecular cluster of SCN9A/SCN10A formed in DRG neurons of mice with diabetic neuropathic pain induced by STZ. (A) A schematic view of the generation of chronic diabetic neuropathy mouse model. (B) Fasting blood glucose concentration of the mice were tested before STZ treatment and 2 weeks after last injection of STZ. (C) The mechanical threshold of mice along the experimental course. (D) Immunostaining of SCN9A (green) and SCN10A (red) showed that supramolecular clusters of SCN9A/SCN10A were detected in DRG neurons from mice with 10-weeks diabetic neuropathy (d-f), not in DRG neurons of naïve mice (a-c), Scale bar=20 μm.

## Discussion

We here show that the percentage of SCN9A/SCN10A neurons increased in DRG of SNI mice at 2 WPO (Fig. 1 B, F), and that Nav1.7/Nav1.8/TrkB formed clusters at 2 WPO and further developed into SMACs through orderly constructing and growing by 6 WPO (Fig. 2). Similar in mice, BPA patients with severe chronic neuropathic pain also had an increase number of SCN9A/SCN10A/TRKB neurons and dramatic upregulated protein expression of SCN9A and SCN10A in TRKB neurons in injured DRG (Fig. 3, 4, 6). SMACs of Nav1.7/Nav1.8/TRKB formed on and along plasma membrane with tens even hundreds of particles constituted with Nav1.7/Nav1.8/TRKB and clusters of Nav1.7/Nav1.8 also formed in nerve of DRG from BPA patients (Fig. 5). We further found that STZ-induced diabetic mice with chronic neuropathic pain also had SMACs of Nav1.7/Nav1.8 in DRG neurons (Fig. 7). These data suggest that excessive expression of SCN9A, SCN10A and TRKB (Fig. 5, 6) could contribute to the formation of SMACs of Nav1.7/Nav1.8/TRKB.

Clustering of membrane proteins plays critical roles in a variety of biologic events, such as signal transduction, asymmetric differentiation, immune response, endocytosis, and etc. For examples, membrane-anchored Ephrin needs be clustered to bring loosely distributed Eph receptors together to induce efficient downstream signaling (*30*). Notch clusters formed in asymmetric divided Drosophila sensory organ precursor cells reinforce Notch signaling by concentrating ligands and receptors to drive cell differentiation and fate commitment (*31*). The quickly formed SMACs containing CD3, CD45, cytoskeletal protein talin, protein kinase C (PKC)-θ and the Src-family kinase Lck and Fyn efficiently triggered activation of T cells by antigen-presenting cells within 30 minutes under the incubation of antigen, and orchestrated signaling in SMACs is required for productive T cell responses (*32, 33*). Moreover, clustered receptors on membrane can amplify the downstream signal induced by same amount of ligands (*34, 35*). It’s also detected in detail by nanoscopy that supramolecular cluster of Syntaxin was composed of 75 densely crowded Syntaxins molecules and exhibited diameter of 50 to 60 nanometers (*36*). The formation of receptor cluster can boost signaling amplitude by only changing the size and geometric structure of clusters (*37*), so *Escherichia coli* controls the size of the clustering of chemotactic receptors on cell surface to detect and respond to wide range concentration (from less than 5 nM to 5 mM) of aspartate (*38*).

Clustering of ion channels also play vital roles in the cellular signals triggering in both muscle and neurons and firing properties-determining. For examples, cluster of voltage-gated Cav1.2 could result in the amplification of Ca^2+^ influx into cardiac muscle and is required to ensure that Ca^2+^ sparks are triggered with a probability of approximately unity (*39, 40*). In similar, cluster of short isoform of Cav1.3 facilitated Ca^2+^ influx into hippocampal neurons (*41*). By using Super-resolution scanning patch-clamp Bhargava A et al. further detected that the cluster of Nav1.5 in crest of cardiomyocytes produced macroscopic currents which was 29-47 folds higher than the single sodium channel in T-tubule area of cardiomyocytes did (*42*). It’s also been proved that cluster of Nav1.1 and cluster of Nav 1.2 formed in prenode can accelerate the speed of axonal conduction before myelination (*43*), and that cluster of Nav1.6 which replace Nav1.1 or Nav1.2 in node of Ranvier in late development (after myelination) determined repetitive firing capability of retinal ganglion and cerebellar Purkinje neurons (*44, 45*).

Therefore, the orderly organized SMACs of Nav1.7/Nav1.8/TRKB in pathological DRG could function in a coordinated manner to contribute to hyperexcitability and severe neuropathic pain through increasing the sensitivity to external and internal stimuli, and the downstream signals produced by SMACs of Nav1.7/Nav1.8/TRKB in the pathological DRG neurons could be enhanced several times than that in the same type of normal DRG neurons.

The decreased population of SCN9A/SCN10A neurons in L4 DRG at 6 WPO and in L5 DRG from 2 WPO to 6 WPO could be due to the increase of neuronal death in injured DRG, because it’s reported that apoptosis increased in DRG of mice with sciatic nerve transection (*46*).

## Conclusion

We here show that nerve injury induced upregulation of Nav1.7 and Nav1.8 expression and formation of SMACs of Nav1.7/Nav1.8/TRKB in DRG neurons in SNI and diabetic mice as well as BPA patients with severe chronic neuropathic pain. These findings suggest that Nav1.7 and Nav1.8 could function in coordination in the orderly organized SMACs to increase the hyperexcitability of DRG neurons, and therefore contribute to severe chronic pain. The cluster of Nav1.7/Nav1.8/TRKB might be the therapeutic target for neuropathic pain, and blocking both Nav1.7 and Nav1.8 could be required to achieve the efficient relief of neuropathic pain.

## Data Availability

All data produced in the present study are available upon reasonable request to the authors.

## Acknowledgements

We thank Prof. Patrik Ernfors for precious suggestions for this project. Changgeng Peng is supported by the National Natural Science Foundation of China (32070977, 51971236, 31871063), and National Major Science and Technology Projects of China (2018ZX09733001-006-005). Liting Sun is supported by the National Natural Science Foundation of China (82101320). Rougang Xie is supported by the National Natural Science Foundation of China (82171212, 81870867) and Ministry of Science and Technology of China (2021ZD0203205). Hongyan Shuai is supported by the National Natural Science Foundation of China (82060149).

## Conflict of interest statement

The authors declare no competing financial interests.

## Notes

### Competing Interest Statement

The authors have declared no competing interest.

### Author Declarations

The avulsed roots of the affected brachial plexus and the dislocated cervical DRG tissues were resected during the surgical process of nerve reconstruction or transplantation under the ethic permit (KY20222228) approved by the Ethics Committee of Xijing Hospital, the first affiliated hospital of Fourth Military Medical University. Normal cervical DRG tissues from naturally aborted 29-weeks human embryos were collected according to the ethic permit (MECDU-201909-1) approved by Dali University.

