## Supplemental Figure 1 for "Nav1.7 and Nav1.8 form supramolecular active clusters with TRKB in mouse and human DRG neurons during development of neuropathic pain"

### Supplementary figure and figure legend

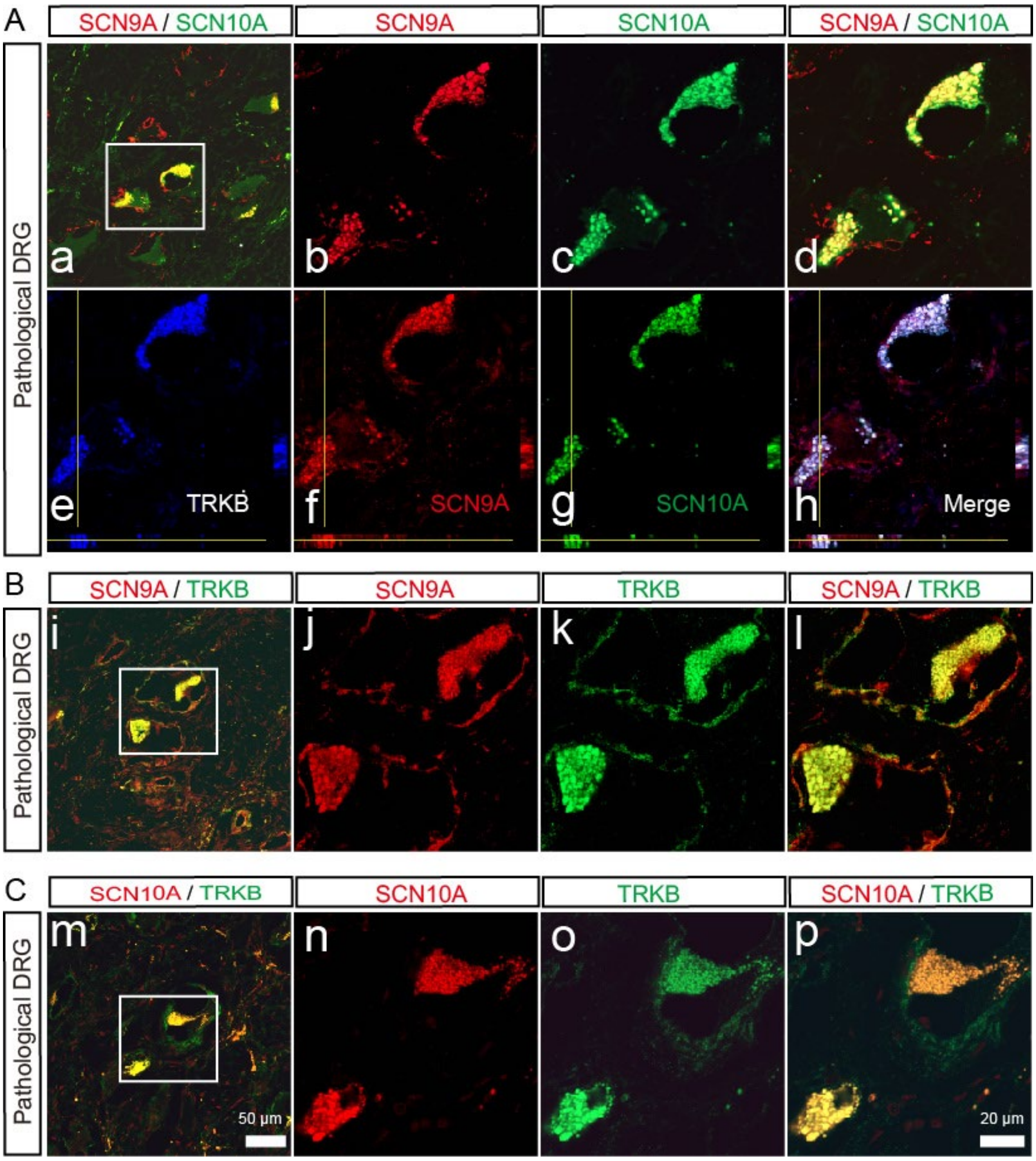


**Fig. S1. Supramolecular cluster of SCN9A/SCN10A/TRKB developed in DRG neurons from BPA patients with NP.** (A) Immunostaining for SCN9A (red), SCN10A (green), and TrkB (blue) showed that they were collocolized in the supramolecular cluster of DRG neurons from BPA patients, b-d are high magnification view of the inset in a, e-h are the projection view of z-stack image of the inset in a. (B) Immunostaining for SCN9A (red) and TrkB (green) showed that supramolecular cluster of SCN9A/TRKB formed on and along plasma membrane of DRG neurons from BPA patients (i-l). (C) Immunostaining for SCN10A (red) and TrkB (green) showed that supramolecular cluster of SCN10A/TRKB formed on and along plasma membrane of DRG neurons from BPA patients (m-p). Scale bar=50 μm (a, i, m) or 20μm.
